# Connectivity changes following transcranial alternating current stimulation at 5-Hz: an EEG study

**DOI:** 10.1101/2023.10.13.23297027

**Authors:** Tien-Wen Lee, Chiang-Shan R. Li, Gerald Tramontano

**Affiliations:** The NeuroCognitive Institute (NCI) Clinical Research Foundation, NJ 07856, US; Department of Psychiatry, Yale University School of Medicine, New Haven, CT 06520; Department of Neuroscience, Yale University School of Medicine, New Haven, CT 06520; Wu Tsai Institute, Yale University, New Haven, CT 06520

**Keywords:** Transcranial alternating current stimulation (tACS), Transcranial electrical stimulation (tES), Electroencephalography (EEG), Exact low-resolution electromagnetic tomography (eLORETA), Coherence, Phase Synchronization, Connectivity

## Abstract

**Introduction:** Transcranial alternating current stimulation (tACS) at 5-Hz to the right hemisphere can alleviate anxiety symptoms. This study aimed to explore the connectivity changes following the treatment.

**Methods:** We collected electroencephalography (EEG) data from 24 participants with anxiety disorders before and after the tACS treatment during a single session. Electric stimulation was applied over the right hemisphere, with 1.0 mA at F4, 1.0 mA at P4, and 2.0 mA at T8, following the 10-10 EEG convention. With eLORETA, the scalp signals were transformed into the cortex’s current source density. We assessed the connectivity changes at theta frequency between the centers of Brodmann area (BA) 6/8 (frontal), BA 39/40 (parietal), and BA 21 (middle temporal). Functional connectivity was indicated by lagged coherences and lagged phase synchronization. Paired t-tests were used to quantify the differences statistically.

**Results:** We observed enhanced lagged phase synchronization at theta frequency between the frontal and parietal regions and between the parietal and temporal regions.

**Conclusion:** Applying tACS 5-Hz over the right hemisphere enhanced inter-regional interaction, which was spectrum-specific and mainly mediated by phase, rather than power, synchrony. The potential neural mechanisms are discussed.

## Introduction

Transcranial alternating current stimulation (tACS) can influence cortical rhythm and activity [1, 2]. Recently, tACS has been proposed as an anxiolytic option [3]. Lee et al. reported that 5-Hz tACS at 2.0 mA over the right hemisphere with the currents oscillating between T8 and F4/P4 (10-10 EEG convention) could alleviate anxiety symptoms effectively [4], called tripod design. It was found that the therapeutic effects were associated with an increase in alpha power and a decrease in beta and gamma power [5], concordant with previous literature about neural markers of anxiety reduction [6–11].

Despite the consistency in the clinical and electrophysiological profiles, enhanced theta power was predicted based on the popular entrainment theory but was not observed [12, 13]. Instead, as stated above, the influence of tACS affected broad spectra (spectrum-unspecific), and accounting for the holistic picture of the neural consequences of tACS required the collaboration of several imperative network mechanisms [5]. To supplement previous findings that focused on regional powers, the primary purpose of this study was to investigate the functional connectivity changes to 5-Hz tripod tACS between the frontal, parietal, and temporal regions.

The effect of tACS was built upon its influences on neuronal activities, i.e., changes in firing rate and spike timing [14–16]. A recent invasive primate study suggested that tACS may influence the timing, not the rate, of spiking activity within the targeted brain region [17]. Such an effect is frequency- and location-specific. Administering in-phase alternating currents at two separate brain regions might synchronize their activities and enhance their connection through neural plasticity [18, 19]. We thus postulated that the connectivity strength between the frontal, parietal, and temporal areas would increase after tACS, and the increment was spectrum specific.

To fulfill the brain-based approach, exact low-resolution brain electromagnetic tomography (eLORETA) was exploited to transform EEG data from the scalp to the gray matter voxels of a template brain [20, 21]. The relationships between the current source density (CSD) time series of different brain regions at distinct spectra were evaluated by lagged coherence and phase synchronization, with the former a linear and the latter a nonlinear index of functional connectivity [22].

## Materials and Methods

### Participants

We reviewed the data collected from our clinics between 2018 and 2022 after obtaining approval from a private Review Board (Pearl IRB; https://www.pearlirb.com/). Twenty-four anxiety patients who received 5-Hz tACS treatment over the right hemisphere and had pre- and post-treatment EEGs were identified. As for detailed clinical profiles and the methodology of tACS, please refer to other reports [4, 5].

### tACS, EEG recording, and pre-processing

The tACS montage of electrodes covered the right lateral side of the head at F4, T8, and P4 positions in terms of the 10-10 EEG convention. The peak current intensity for T4 was 2.0 mA, while those at F4 and P4 were 1.0 mA. Alternating sinewave currents oscillated at 5 Hz between electrodes T4 and F4/P4 for 25 minutes. Before and after the tACS treatment, we used the Brainmaster device (https://brainmaster.com/) to acquire 10 min eye-open digital EEG data at 256 samples/sec with linked-ear reference.

The EEG traces were edited using the Software EEGLAB [23]. A band-pass filter (1-50 Hz) was applied to preprocess the data, followed by automatic artifact removal (Artifact Subspace Reconstruction). A single rater (TW Lee) manually eliminated any remaining noisy portions. The clean EEG data, which included the removal of various artifacts such as blinks and eye movements, were then segmented into 2-second epochs and imported into eLORETA for further analyses.

### eLORETA analyses

The eLORETA is a tomographic method for deriving electric neuronal activity from EEG by computing a weighted minimum norm inverse solution, where the weights are adaptive to the data (data-dependent) and the formula may accommodate measurement and biological noises [20, 21]. Any arbitrary point-test sources can be correctly computed with exact, zero-error localization. eLORETA utilizes the principles of linearity and superposition to effectively identify distributed electric sources (i.e., CSD) within the brain cortex (a template with 6,239 gray matter voxels), although its spatial resolution may be limited. The CSD time series were decomposed to those of the following power spectrum, delta (1–4 Hz), theta (4–8 Hz; the focus of this study), alpha (8–12 Hz), low beta (12-15 Hz), mid-to-high beta (15–30 Hz), and low gamma (30–45 Hz).

### Connectivity and statistical analyses

Lagged (general) coherence and phase synchronization were adopted to represent linear and nonlinear functional connectivity strengths, respectively [22]. Both were derived from the CSD time series of the selected voxel. The equations to obtain the latter are the same as the former except for a pre-normalization step to discount the influence of power, hence non-linear. Namely, the relationship between amplitudes did not affect phase synchronization. Total coherence is the sum of the lagged and instantaneous dependence, and the latter contains confounds from the “non-physiological contribution due to volume conduction and low spatial resolution” [22]. Accordingly, the instantaneous part was disregarded in this report (the same reason for the lagged phase synchronization).

The average connectivity strengths were calculated between three coordinates, which were the center of Brodmann area (BA) 6/8 (frontal; [25,0,50]), BA 39/40 (parietal; [45,-50,35]), and BA 21 (middle temporal; [60,-15,-15]). Paired t-tests were used to examine the connectivity changes before and after the tACS (3 connections in total).

## Results

The mean age of the 24 selected patients was 34.6 (SD 14.2), ranging from 15.2 to 54. The gender ratio male to female was 9:15. Our hypothesis was verified that the connectivity strengths were increased after the 5-Hz tACS tripod treatment. The enhancement was statistically significant for phase synchronization after multiple comparison corrections. It was noted in the connections between the frontal and parietal regions and between the parietal and temporal areas, not between the frontal and temporal counterparts. The connectivity strengths and the statistics are summarized in Table 1, and the significant results are illustrated in Figure 1. Supplementary analyses showed that the connectivity changes were only present in the theta range, not in other spectra (spectrum-specific; data not shown).

**Table 1.**
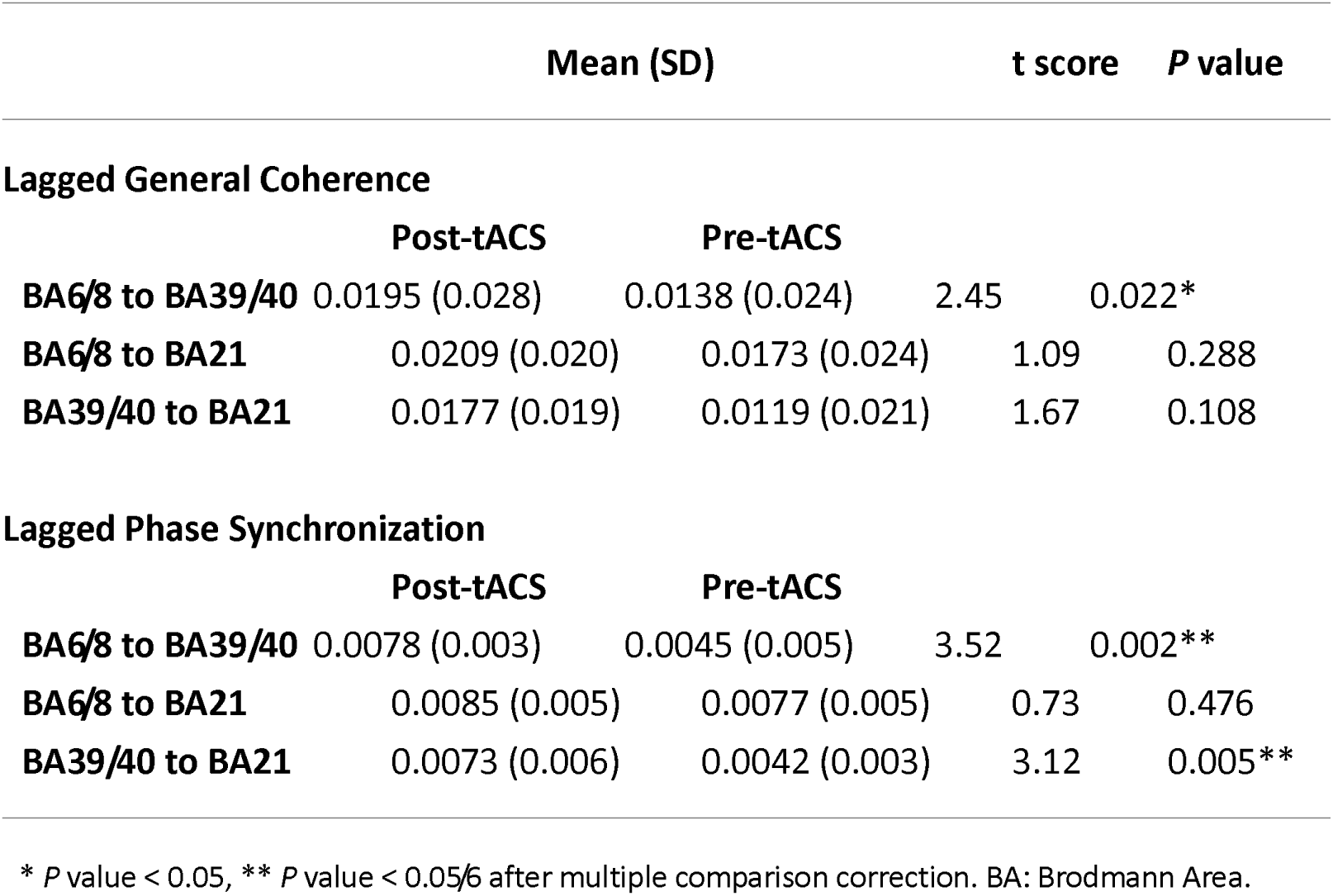
Lagged general coherence and phase synchronization were computed between the central coordinates of BA 6/8, BA 39/40, and BA 21 and were compared between post-tACS and pre-tACS conditions.

**Fig. 1.**
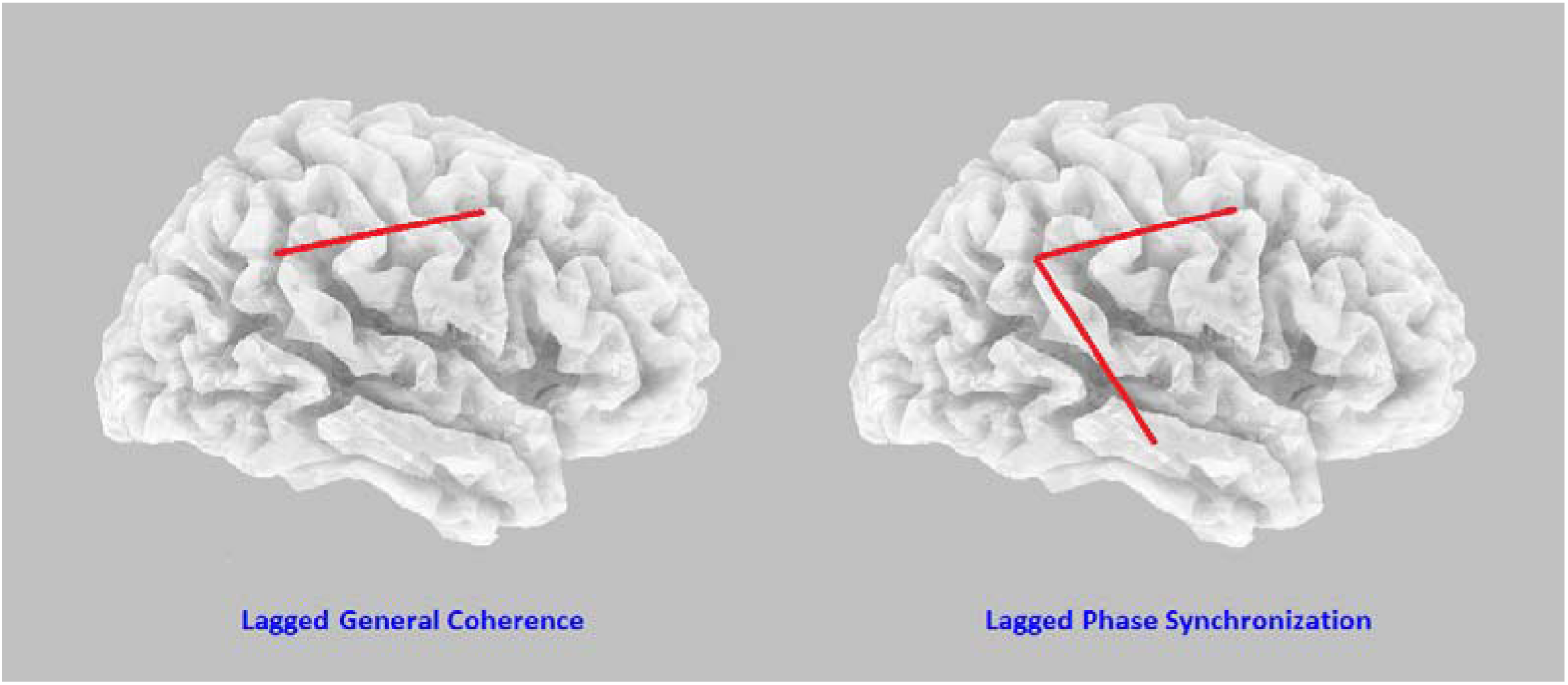
Left: The lagged coherence was increased in the frontoparietal network. Right: The lagged phase synchronization was increased in both the frontoparietal network and parietal-temporal connection.

## Discussion

This research studied the connectivity strength changes after tACS at 5-Hz and 2.0 mA for 25 minutes, with a tripod montage covering the frontal, parietal, and temporal regions. The design was proposed as a quick way to ease anxiety [4]. Pre- and post-treatment EEGs were collected and analyzed for 24 participants using an imaging method eLORETA, which converted the signals recorded at the scalp to CSD in the brain cortex. Average functional connectivity was thus derived from the CSD time series between the frontal, parietal, and temporal regions. After tACS treatment, the lagged phase synchronization at theta range (i.e., spectrum-specific) significantly increased in the frontal–parietal and the parietal–temporal connections. The lagged coherence was enhanced between the frontal and parietal regions with a *P* value lower than 0.05, although not surviving Bonferroni correction. Our hypothesis was generally verified but with some caveats, discussed below.

Strong evidence suggested that tACS may influence the timing of neuronal spikes [14, 15, 17]. When tACS is delivered to two distinct brain regions, the firing of neural tissues in these regions is expected to synchronize more closely by the applied frequency. Furthermore, given that the recorded EEGs were not in real-time with the delivered tACS, the significant inter-regional interactions indicated the engagement of a plasticity mechanism. Interestingly, the index derived from lagged phase synchronization was more robust than that of lagged coherence. Our previous study investigating the regional power changes to tACS demonstrated complicated spectral power features incompatible with entrainment theory [5]. From a mathematical perspective, the formula of phase synchronization is similar to that of coherence, except for a normalization procedure to discount the influence of the power. Based on the two analytic results (power and connectivity), it was deduced that the inter-regional modulatory effects of tACS were not due to the concurrent entrained oscillation/power at multiple regions. Rather, it was through synchronizing their phase relationship, echoing previous neurophysiological research that recorded and investigated spike timing under tACS [17]. Compatible with our conjecture, Vossen et al. explored the aftereffects of alpha tACS and concluded that the modulatory effect of tACS was mediated by plasticity rather than entrainment [24]. In summary, at the large-scale network level, the inter-regional influence of tACS was mediated by synchronization in phase (crosstalk), not by the concurrent entrainment of powers (regional profile).

It was noticed that the connectivity changes were not significant between the frontal and temporal regions. Again, if the influence of tACS on neural connectivity mainly worked through concurrent entrainment across targeted areas, the interactions between the three explored regions would tighten altogether. We inferred that the differential manifestations originated from the discrepancy in the hardwire underpinnings. It was noted that the superior longitudinal fasciculus (SLF) bridges between the frontal and parietal regions and between the inferior parietal cortex (BA39/40) and the middle temporal cortex (BA 21) (SLF III). The former constitutes the frontoparietal network, and the latter links the two cortical nodes of the default-mode network [25]. No white matter “highway” exists between the dorsolateral prefrontal and middle temporal cortices (note: the inferior frontal cortex and anterior temporal region are connected by uncinate fasciculus). It hints at one of the most paramount plasticity mechanisms, spike-time-dependent plasticity, which requires direct axonal connections to take effect [18, 19]. The lack of significant interaction between the dorsolateral prefrontal cortex and middle temporal cortex supports a plasticity mechanism of tACS, arguing against the entrainment theory.

It was observed that applying a particular frequency of tACS can “entrain” or “synchronize” the neural oscillations to match the frequency of the electrical stimulation [12, 13], framed as an entrainment theory. However, our recent report demonstrated that narrow band 5-Hz tACS desynchronized neural oscillation, which affected broad spectra beyond the default frequency of tACS [5]. An earlier study by Brignani et al. challenged the idea that tACS effectively modulated brain oscillations [26]. Alexander et al. showed that 10-Hz tACS, in fact, reduced alpha power in the frontal region [27]. In addition, Lafton et al. applied the intracranial recording and observed no sleep rhythm entrainment to tACS [28]. The contradictory findings cannot be resolved by entrainment theory alone but require a broader mechanism to reconcile them. Agreeing with Vossen et al. [24], we believe that spectrum- and region-specific neural plasticity could be a better candidate to accommodate the tACS influence on regional powers and inter-regional interactions. Nevertheless, we cannot exclude the possibility that injecting an artificial narrow-band alternating current may interfere with underlying neural synchronization under certain conditions, given our previous analysis and several other reports summarized above [5]. It is noteworthy that even if tACS impedes the underlying neural synchronization, it may still be beneficial. In our previous report, power reduction in the right hemisphere due to tACS might reduce emotion reactivity according to emotion lateralization theory and hence, might catalyze the anxiolytic effect [5, 29]. The merits and demerits of tACS thus could be context-dependent, which requires further research to clarify.

## Conclusion

The neural influence of tACS is still under active investigation. This research explored the connectivity changes following 5-Hz tACS over the right hemisphere. Increased lagged phase synchronization at theta spectrum was noticed between frontal and parietal regions and between parietal and temporal regions. The enhancement in functional connectivity was likely mediated by its influence on neural spike timing and spike-time-dependent plasticity.

## Authors Contributions

All authors contributed intellectually to this work. TW Lee carried out the analysis and wrote the first draft. All authors revised and approved the final version of the manuscript.

## Data Availability

All data produced in the present study are available upon reasonable request to the authors

## Acknowledgments

This work was supported by NeuroCognitive Institute (NCI) and NCI Clinical Research Foundation Inc. We would like to thank Almeida Sergio for helping prepare the research material.

## Statements and Declarations

All authors declare no conflicts of interest.

## Compliance with ethical standards

This research analyzed the databank collected from 2018 to 2022. The authors assert that all procedures contributing to this work comply with the ethical standards of the relevant national and institutional committees on human experimentation and with the Helsinki Declaration of 1975, as revised in 2008.

